# Predictors of Mental Health during the Covid-19 Pandemic in the US: role of economic concerns, health worries and social distancing

**DOI:** 10.1101/2020.06.06.20124198

**Authors:** Fabrice Kämpfen, Iliana V. Kohler, Alberto Ciancio, Wändi Bruine de Bruin, Jürgen Maurer, Hans-Peter Kohler

## Abstract

**Objective:** To assess mental health in the US adult population in the Covid-19 pandemic and explore the roles of economic concerns, health worries and social distancing in shaping mental health outcomes.

**Methods:** We analyze online survey data from the “Understanding America Study” (UAS) that is representative of the US adult population and covers the period of March 10-31st 2020 (sample size: 6436).

**Results:** About 29% (CI:27.4-.30.4%) of the US adult population reported some depression/anxiety symptoms osver the study period, with symptoms deteriorating over the month of March. Worsening mental health was most strongly associated with concerns about the economic consequences of the pandemic, while concerns about the potential impact of the virus on respondents’ own health and the practice of social distancing also predicted the presence of depression and anxiety symptoms, albeit less strongly.

**Conclusions:** Our findings point towards a major mental health crisis unfolding simultaneously with the pandemic in the US. They also highlight the importance of economic countermeasures and social policy for mitigating the impact of Covid-19 on adult mental health in the US over and above an effective public health response.

## 1. Introduction

Among the myriad of consequences that the Covid-19 pandemic has on the health care, economic and social spheres in the US and worldwide, experts and policy makers are increasingly urging to consider the mental health consequences of the pandemic.^[1,2]^ Some observers even went as far as calling mental illness resulting from Covid-19 the “inevitable” next pandemic.^[3]^ The importance and urgency to address the short and long term aftermaths of Covid-19 on individual and population level mental health have been outlined in a recent policy brief issued by the United Nations.^[4]^ Despite this compelling need to generate knowledge about the Covid-19 impact on mental health of individuals and populations, research on the mental health consequences of the pandemic, especially in the US, is still sparse and the determinants and sociodemographic patterns of mental well-being during Covid-19 are still not well documented. Prior evidence suggests that the experience of large scale disasters is associated with increases in depression and anxiety, post-traumatic stress disorder (PTD), and a broad range of other mental disorders ^[1,5,6]^. In the context of Covid-19, these mental health implications may be amplified by factors such as increased uncertainty related to individual’s own health because of the exposure to a new highly infectious disease ^[6–8]^, the profound economic consequences of the pandemic in the US and globally, the implementation of lock-down measures ^[9–12]^, resulting in the practice of prolonged social distancing. ^[1,13–16]^

We investigate the consequences of the Covid-19 pandemic on mental health in the US between March 10th and March 31st, 2020, the early period of the pandemic when many US residents began to realize that Covid-19 was about to fundamentally affect their lives.^[9]^ Using nationally representative population-based data for the US adult population from the *Understanding America Study*, ^[17]^ we focus on three potential pathways through which the pandemic can affect mental health: uncertainties and perceptions related to the immediate impact of the coronavirus on own health, concerns about the economic consequences related to the pandemic and impact of practicing social distancing.

## 2. Methods

### Understanding America Study (UAS)

Our analysis utilizes the Covid-19 focused questionnaire of the UAS,^[17]^ implemented between March 10-31, 2020. UAS is a nationally representative probability-based Internet panel of approximately 8,500 respondents administered in English and Spanish. As part of its address-based study recruitment, UAS provides Internet access and a tablet to all panel members who may otherwise not be able to participate in the study.^[18]^ In our study sample, 4.4% of the respondents were provided with an Internet-connected tablet at the time of recruitment in order to address the “digital divide” between different population groups (Table A1 in Online Appendix). A comparison of UAS data with data from the *Current Population Survey* (CPS) and the *Health and Retirement Study* (HRS) shows that UAS lines up well with the CPS on a number of common variables, and matches the quality of the HRS, a traditional survey considered as the gold standard in social research.^[19,20]^ See Online Appendix 1 for more details on UAS.

By March 31st 2020, out of the 8,815 participants to whom the questionnaire was fielded, 6,885 individuals (78.1%) completed the survey. There were no significant differences in terms of age, gender and education between individuals who did and who did not complete the survey (Online Appendix Table A2), even if non-completion was significantly higher among married UAS participants (*χ*^2^ = 5.659, p-value= 0.017).

### Measurement of mental health in UAS

Mental health, i.e., presence of depressive and anxiety symptoms, is assessed by the Patient Health Questionnaire-4 (PhQ-4) which has been validated in the US. ^[21]^ The PhQ-4 is an ultra-brief four-item scale for detecting depression and anxiety, which represent the most common mental disorders during periods of disasters and disease outbreaks ^[22,23]^ and are often co-occurring. ^[24–28]^ Composed by four distinct questions that are answered on a four-point Likert scale ranging from 0 (“Not at all”) to 3 (“Nearly every day”), this scale has internal reliability and construct validity and is a reliable instrument for screening for both depressive and anxiety symptoms outside of clinical settings. ^[21,29]^ (See Online Appendix 2 for more details on PhQ-4).

PhQ-4 scores ranged from 0 to 12, and we categorized respondents as having *no depression/anxiety symptoms* if their score was 0, 1 or 2, *mild depression/anxiety symptoms* if their score ranged from 3 to 5, *moderate depression/anxiety symptoms* if they scored 6 to 8 and *severe depression/anxiety symptoms* if their score was 9 or higher. ^[21]^

Importantly, the UAS provides information on the respondents’ mental health prior to the Covid-19 outbreak, albeit based on a different survey item. Between 2018 and 2020, panel participants were asked whether they agree or not to the statement: “I am someone who is depressed, blue”, with five permissible answers ranging from “strongly disagree” to “agree strongly”. Although not fully comparable with the PhQ-4 scale, we used this information as a baseline measure for the presence of depressive symptoms to control for any underlying differences in mental health characteristics among the UAS respondents prior to the outbreak of Covid-19 in the US.

### Measurement of Covid-19 related risks, behaviors and events

In addition to sociodemographic characteristics such as sex, age, education, race and marital status at the time of the interview (see descriptive statistics in Online Appendix Table A1), UAS elicited respondents’ perceptions that specific Covid-19-related events will occur, using a validated visual linear scale ranging from 0 to 100.[30] The survey asked respondents to rate their probability of getting infected with coronavirus in the next three months and the probability of dying in case of infection. We generated a measure of respondents’ *perceived risk* of excess mortality due to Covid-19 by multiplying these two probabilities. To measure economic concerns about the Covid-19 impact, survey participants were asked to rate the chances that they will run out of money within the next three months. The respective questions can be found in the Online Appendix 2. UAS also asked participants whether they have reduced their social life and activities because of the pandemic. To measure social distancing, we generated a dichotomous variable that takes the value of one if respondents stated that they have canceled or postponed travel for pleasure, and/or canceled or postponed personal or social activities, and/or avoided public space gatherings or crowds and/or eating at a restaurant (Cronbach’s *α*=0.804), indicating good reliability of the items to measure the same construct ^[31]^.

To assess the impact of the increasing caseloads in Covid-19 infections and deaths on mental health in the US, we matched to our UAS sample publicly available daily data from state and local governments, and health departments on the number of cases and deaths in the US during our study period in March.^[32]^ The match was based on the day when the UAS respondents completed the survey.

### Statistical analysis

To explore the pathways through which the Covid-19 pandemic affects mental health in the US, we estimated separate ordered probit regression models with three different explanatory variables that reflect: *1)* individual’s economic concerns induced by the pandemic (measured by the perceived probability of running out of money on a continuous scale); *2)* distress due to the immediate health impacts of the virus (measured by the probability of own Covid-19 excess mortality on a continuous scale); and *3)* the influence of practiced social distancing. The outcome variable in all three models is a categorical variable that indicates the presence and severity of depression/anxiety symptoms measured by the PhQ-4, as defined above, where higher values indicate presence of more severe symptoms.

Usual standardization to compare the strength of these associations with mental health is inadequate because social distancing is measured by a dichotomous variable. Hence, to ensure a more meaningful comparison between the three explanatory variables, we subtracted the mean and re-scaled the two continuous variables by dividing them by *γ* = 2.103 times their respective standard deviation so that the coefficients of the continuous variables correspond in magnitude to a marginal increase in the dichotomous variable from 0 to 1 (see Online Appendix 3 for more details).

In addition to a set of sociodemographic control variables, our models also controlled for state fixed effects, the self-reported baseline depression level and the year and month when this measure was collected. The model specifications also included time fixed effects (in days) to capture the influence of aggregate effects, such as public announcements at the federal level or national news, that are shared among UAS respondents on a given day. Our analysis used post-stratification weights, generated through a raking algorithm, to align the sample to the US adult population in terms of gender, race/ethnicity, age, education and geographic location (see https://uasdata.usc.edu/page/Weights and Angrisani et al. ^[19]^). Analysis was performed with StataSE 14.

## 3. Results

Table 1 summarizes the weighted distribution characteristics of the main variables used in our analysis. The weighted mean of the PhQ-4 score in the US was about 1.9, with 71.1% of respondents reporting no depressive/anxiety symptoms, while 17.8% reported mild depressive/anxiety symptoms; 6.6% of the sample had moderate and 4.6% experienced severe depressive/anxiety symptoms.

**Table 1:** Summary statistics – UAS Sample 230

| Variables | Scale | Pop.<br>mean<br>March 10-31 | Pop.<br>std deviation<br>March 10-31 | Pop.<br>mean<br>March 10-16 | Pop.<br>std deviation<br>March 10-16 |
| --- | --- | --- | --- | --- | --- |
| **Outcome variables** |  |  |  |  |  |
| PhQ-4 score | From 0 to 12 | 1.944 | 2.829 | 1.910 | 2.861 |
| No depression/anxiety symptoms | Yes (1)/no (0), PhQ-4 score 0-2 | 0.711 | 0.453 | 0.723 | 0.448 |
| Mild depression/anxiety symptoms | Yes (1)/no (0), PhQ-4 score 3-5 | 0.178 | 0.382 | 0.165 | 0.371 |
| Moderate depression/anxiety symptoms | Yes (1)/no (0), PhQ-4 score 6-8 | 0.066 | 0.247 | 0.064 | 0.244 |
| Severe depression/anxiety symptoms | Yes (1)/no (0), PhQ-4 score 9+ | 0.046 | 0.209 | 0.049 | 0.215 |
| **Control variables** |  |  |  |  |  |
| Chances to run out of money | Continuous scale from 0 to 100, rescaled (/100) | 0.158 | 0.261 | 0.132 | 0.237 |
| Excess mortality | Continuous scale from 0 to 1 | 0.043 | 0.093 | 0.038 | 0.090 |
| Social distancing | Yes (1)/no (0) | 0.655 | 0.475 | 0.577 | 0.494 |
| **Baseline depression** |  |  |  |  |  |
| I am someone who is depressed, blue | Strongly disagree (1)–Agree strongly (5) | 2.210 | 1.285 |  |  |
| Strongly disagree | Yes (1)/no (0) | 0.438 | 0.496 |  |  |
| Disagree a little | Yes (1)/no (0) | 0.171 | 0.377 |  |  |
| Neither Agree nor Disagree | Yes (1)/no (0) | 0.179 | 0.383 |  |  |
| Agree a little | Yes (1)/no (0) | 0.165 | 0.372 |  |  |
| Agree strongly | Yes (1)/no (0) | 0.046 | 0.210 |  |  |

*Note:* Source of Data: “Understand America Study” (UAS), survey 230, collected between March 10 and March 31, 2020. All statistics use sample weights to make the survey representative of the U.S. population aged 18 and older. Baseline depression level is defined based on the question: “I see myself as someone who is depressed, blue”. This question was asked in UAS survey 121, fielded between Jan 2018 and March 2020. We consider someone as having practised social distancing if a respondent “canceled or postponed travel for pleasure”, “canceled or postponed personal or social activities”, “avoided public spaces, gatherings or crowds” or “avoided eating at restaurant”. “Pop.” is short for population and “std.” is short for standard

During the period March 10-31st, 2020, respondents reported a 15.8% probability to run out of money within three months because of the pandemic’s impact. This is higher than the population mean reported previously for the period March 10–16th (13.2%) using the UAS survey ^[9]^, indicating a deterioration in the perceived economic perspectives in the US during the month of March. Similarly, our sample showed a weighted mean of perceived excess mortality due to Covid-19 of about 4.3%, which is also higher than the previously reported mean (3.8%) based on the UAS.[9] Table 1 also shows that less than two thirds of the US population were practicing social distancing during the study period.

Figure 1 shows weighted means of PhQ-4 score on the left and weighted proportions of the US population that has a least some depressive/anxiety symptoms on the right over the period March 10-31. We use post-stratification weights so that the weighted means and proportions are representative of the US population for ***each particular time period on the x-axis***. Reports of depression/anxiety symptoms increased over time in March, reaching the highest point in the last week of the month. This increase was not driven by persons with higher depression level completing the online survey later in the month, as the trend is not increasing when we consider depression level at baseline instead of the PhQ-4 mental health measure collected in March (see Figure A1 in Online Appendix).

**Figure 1:**
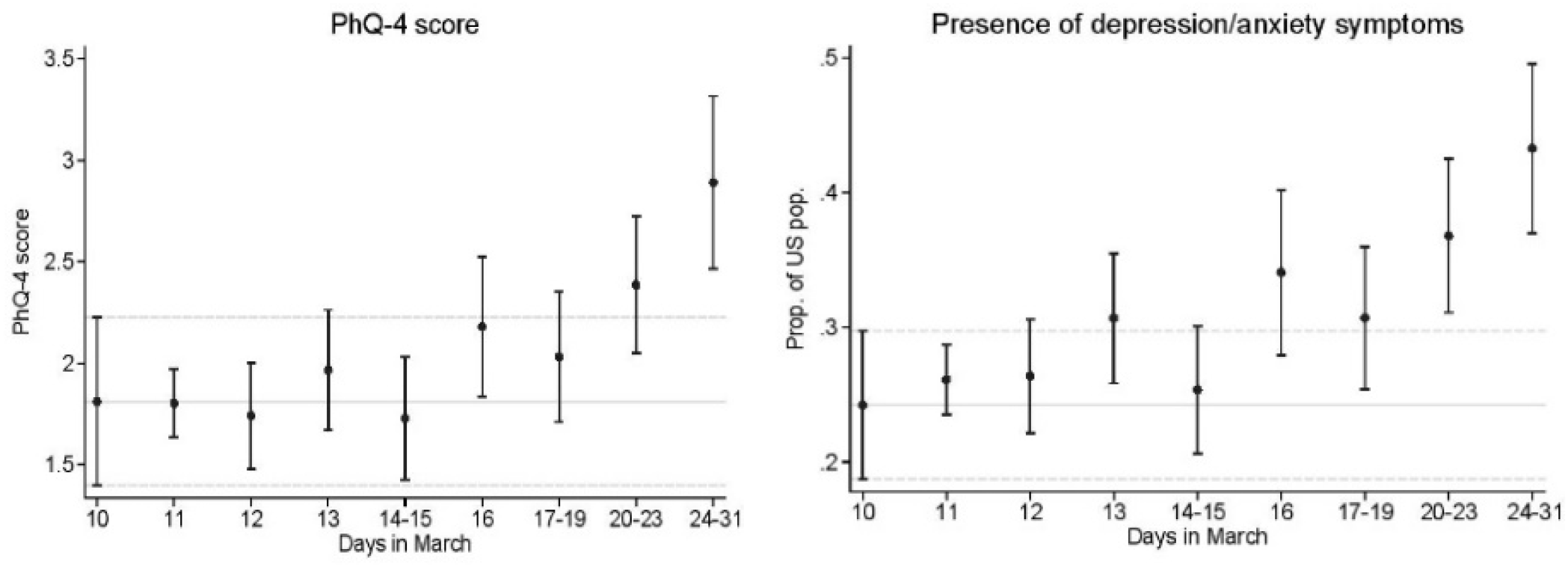
Changes in the mental health (PhQ-4) from March 10th to March 31st *Notes*: Source of Data: “Understanding America Study” (UAS), survey 230, collected between March 10 and March 31, 2020. Plots shows weighted means, along with 95% confidence intervals, of PhQ-4 score on the left and weighted proportions of the US population that has at least some depressive/anxiety symptoms on the right. We use post-stratification weights so that the weighted means and proportions are representative of the US population for ***each particular time period*** on the *x*-axis.

We also investigated the associations between PhQ-4 and the raising number of Covid-19 infection cases and deaths during the period March 10-31, 2020 (Table A3 in the Online Appendix). A one-unit increase in the log number of Covid-19 cases resulted on average in about 0.090 increase in the latent mental health ordered probit index (z-score). This increase corresponded to a drop of 2.6 percentage points in the probability of reporting no depressive/anxiety symptoms, whereas the probability of reporting mild, moderate and sever symptoms increases by 1.2, 0.7 and 0.7, respectively (Online Appendix Table A4). This association was particularly strong for males and college graduates.. Similar patterns were also estimated for the impact of the number of deaths in the US, where a one-unit increase in the log number of Covid-19 deaths led on average to an increase of about 0.104 in the latent mental health index, indicating higher levels of depression/anxiety, with this effect being again stronger for males and college graduates (Table A3 columns 3 and 5 in Online Appendix).

High expected probability of running out of money is positively associated with the PhQ-4 mental health score (Table 2, Column 1). Similarly, perceived excess mortality and social distancing (Columns 2 and 3) are also strongly correlated with higher levels of depression/anxiety. Column 4 shows that when all of the three explanatory variables are included in the same model specification, they remain strongly associated with the PhQ-4 mental health score, “independently” from each other. However, because of the different scales in which these variables are measured, the magnitude of their associations cannot be directly compared. Column 5 allows a direct comparison of the coefficients after standardizing the continuous variables as explained above. The association of the probability of running out of money with mental well-being is stronger than the two other variables capturing alternative pathways affecting mental health during the pandemic. Social distancing shows the second-strongest association, while perceived own excess mortality due to Covid-19 has the weakest association with mental health among the three. The result of a one-sided z-test rejects the null hypothesis that the coefficient associated with social distancing is equal or larger than the coefficient associated with running out of money (p-value= 0.027). Average marginal effects reported in Table A5 show that a *γ* standard deviation increase in the perceived probability of running out of money because of Covid-19 led to a 11.1 percentage points decrease in the probability of reporting no depressive/anxiety symptoms, whereas it increases the probability of showing mild, moderate and severe symptoms by 5.1, 3.0 and 3.1, respectively. The magnitude of the average marginal effects of the two other pathways are about half of those of the economic pathway, with social distancing having larger influences on mental health than perceived health concerns.

**Table 2:** Associations of economic uncertainties, perceived health risks and social distancing with mental health score (PhQ-4)

|  | PhQ-4<br>(1) | PhQ-4<br>(2) | PhQ-4<br>(3) | PhQ-4<br>(4) | PhQ-4<br>rescaled<br>(5) |
| --- | --- | --- | --- | --- | --- |
| Economic uncertainties (i.e. running out of money) | 0.850***<br>[0.672,1.028] |  |  | 0.724***<br>[0.537,0.911] | **0.398\*\*\***<br>**[0.295,0.500]** |
| Perceived health risks (i.e. own excess Covid-19 mortality) |  | 1.336***<br>[0.907,1.766] |  | 0.846***<br>[0.405,1.286] | **0.165\*\*\***<br>**[0.079,0.251]** |
| Social distancing |  |  | 0.307***<br>[0.204,0.410] | 0.241***<br>[0.136,0.347] | **0.241\*\*\***<br>**[0.136,0.347]** |
| Observations | 6436 | 6377 | 6434 | 6368 | **6368** |

*Notes:* Results of weighted ordered probit regressions with robust confidence intervals in brackets, \* $p < 0.1$ , \*\* $p < 0.05$ , \*\*\* $p < 0.01$ . Specifications include state and time fixed effects. The list of control variable includes: sex, age and age<sup>2</sup>, educational level (binary variable for each category), race and whether the respondent was married at the time of the interview. We control for depression level at baseline, along with the year and month when this measure was collected. We use sample weights to make the survey representative of the U.S. population aged 18 and older. Data come from “Understanding America Study” (UAS) collected between March 10 and March 31, 2020. “Running out of money” and “Excess mortality” in Column 5 have been standardized by subtracting their respective mean and dividing by $\gamma \times$ their standard deviations.

Table 3 reveals important heterogeneity in these patterns, with perceived economic uncertainty being particularly important for males (Columns 1 and 2), whereas concerns about own health being less important for males and non-while individuals (Columns 1,2, 5 and 6). The influence of social distancing on PhQ-4 mental health score however appears to be relatively similar across the various sociodemographic groups. Results including the interaction terms between our three main independent variables and the various sociodemographic groups are presented in Table A6 in the Online Appendix.

**Table 3:** Differences in the associations of economic uncertainties, perceived health risks and social distancing with mental health score (PhQ-4)

|  | PhQ-4<br>Female | PhQ-4<br>Male | Difference<br>Male vs<br>Female<br>p-value | PhQ-4<br>No college | PhQ-4<br>College | Difference<br>No college vs<br>College<br>p-value | PhQ-4<br>White | PhQ-4<br>Non-white | Difference<br>White vs<br>Non-white<br>p-value |
| --- | --- | --- | --- | --- | --- | --- | --- | --- | --- |
|  | (1) | (2) |  | (3) | (4) |  | (5) | (6) |  |
| Economic Uncertainties<br>(i.e. running out of<br>money) (std) | 0.311***<br>[0.186,0.435] | 0.541***<br>[0.380,0.701] | 0.021 | 0.416***<br>[0.296,0.535] | 0.371***<br>[0.169,0.573] | 0.705 | 0.367***<br>[0.256,0.478] | 0.466***<br>[0.252,0.680] | 0.410 |
| Perceived health risks<br>(i.e. own excess Covid-19<br>mortality) (std) | 0.222***<br>[0.120,0.325] | 0.073<br>[-0.070,0.216] | 0.090 | 0.165***<br>[0.064,0.266] | 0.164**<br>[0.018,0.309] | 0.988 | 0.217***<br>[0.121,0.312] | -0.007<br>[-0.199,0.186] | 0.040 |
| Social distancing | 0.212***<br>[0.078,0.346] | 0.267***<br>[0.105,0.429] | 0.600 | 0.202***<br>[0.070,0.334] | 0.338***<br>[0.178,0.498] | 0.190 | 0.237***<br>[0.125,0.350] | 0.264**<br>[-0.000,0.528] | 0.854 |

*Notes:* Results of weighted ordered probit regressions with robust confidence intervals in bracket, \* $p < 0.1$ , \*\* $p < 0.05$ , \*\*\* $p < 0.01$ . Specifications include state and time fixed effects. The list of control variable includes: sex, age and age<sup>2</sup>, educational level (binary variable for each category), race and whether the respondent was married at the time of the interview. We control for depression level at baseline, along with the year and month when this measure was collected. We use sample weights to make the survey representative of the U.S. population aged 18 and older. Data come from “Understanding America Study” (UAS) collected between March 10 and March 31, 2020. “Running out of money” and “Excess mortality” have been standardized by subtracting their respective mean and dividing by $\gamma \times$ their standard deviations.

## 4. Discussion

We assessed factors associated with mental health of US adults age 18+ years during the early outbreak of the Covid-19 pandemic, March 10-31, 2020. During this period, the virus spread rapidly in the US, with confirmed infection cases and deaths rising exponentially, from 1018 to 187,962 and from 31 to 3630, respectively.^[32]^ We used the clinically validated PhQ-4 instrument to assess the presence of depressive and anxiety symptoms in a nationally representative sample of US adults aged 18+ years. To our best knowledge, our study is one of the first to document the decline in US adults’ mental health during the early period of the Covid-19 pandemic.

In March, symptoms of depression and anxiety among US adults were increasing and this worsening of mental health was associated with the increasing number of confirmed Covid-19 cases and related deaths in the US. However, increased caseloads was not the primary driving force behind this deterioration of mental well-being among US adults. Rather, concerns about the economic consequences of the pandemic in the near future (i.e., three months from the date of the interview) emerged as the strongest predictor of declining mental health. Worries about the potential impacts of Covid-19 on own health (i.e., risk of infection and as result increased risk of death), as well as the practice of social distancing played relatively smaller roles in predicting poor mental well-being during the early stages of the pandemic. We also documented significant gender differences in the associations of mental health with perceived economic and health risks: Economic concerns are more strongly associated with worse mental health in men than women, whereas the association of mental health with concerns related to own health outcomes is stronger in women than men, which may partly reflect differences in gender roles and behaviors. ^[33–35]^

Moreover, we only find a positive association between perceived risk of Covid-19-related excess mortality and mental health decline for white respondents, with no such association among non-whites. These ethnic/racial differences may be related to corresponding differences in the socioeconomic impact of Covid-19 across racial/ethnic groups with economic concern/worries being a main driver of depression among minorities. ^[36]^

The strong evidence we provide on the short-term implications of Covid-19 on mental health emphasizes the importance of future research on this topic and specifically the need to investigate the long-term effects of the pandemic on mental health outcomes. For instance, prolonged social distancing measures may result in stronger impact on mental well-being at later stages of the pandemic compared to the weak relationship established in this study. With Covid-19 tests becoming increasingly available, the positive outlooks for the developments of potential treatments and vaccines, and positive news coming from countries that were able to contain the spread of the pandemic might however attenuate the health concerns associated with mental health. The long-term economic concerns and their implications on mental health are of particular research interest especially given the dramatic increases in unemployment in the US that have occurred after the data collection for this study was completed. Given the relatively weak social safety net in the US compared to European high-income countries, the importance and urgency to address the aftermaths of Covid-19 on individual and population level mental health is and will be all the more critical to address in the US.

### Limitations

Our analysis data did not allow to firmly establish causality. Nevertheless, our findings are robust to adjustments for respondents’ past mental health history that could potentially confound our results. While our sample is representative for the US adult population, our estimates did not include minors below age 18, for whom economic concerns may not be at the forefront, but whose mental well-being maybe as well affected by the disruption of their daily routines, schooling, extra-curriculum activities, exposure to stress in the household and increased domestic abuse and violence.

PhQ-4 may capture only probable depression/anxiety or psychological distress in response to the abnormality of the Covid-19 pandemic and its implications as opposed to clinical diagnostic, which does not reduce the importance of our findings for public health policy. Although most adults report only mild to moderate levels of depression and anxiety, these may nonetheless affect many important outcome such as work productivity and savings ^[37,38]^

## 5. Public health implications

Our evidence on the short-term implications of the early Covid-19 pandemic on mental health highlights the importance of imminent mental health challenges of the pandemic among US adults, which may further increase as Covid-19 continues to unfold. As a result, there appears to be an increasing need for prevention and mental health services as a consequence of the pandemic, requiring a parallel strengthening of such efforts during the pandemic. Policy makers, health care providers and social workers should therefore plan for a substantial increase in service needs since the ramifications of the Covid-19 pandemic will be likely felt for an extended period of time beyond getting the virus under control and the curve flattened. In addition, our findings highlight the major importance of economic considerations for US adults’ mental health early in the pandemic over and above the evident health concerns and challenges associated with social distancing. Our results suggest a considerable role for economic countermeasures and social policy for mitigating the economic impacts of the Covid-19 pandemic on US adults’ livelihoods and, thereby, helping to protect their mental health and well-being through this unfolding pandemic.

## Online Appendix

### 1. Understanding America Study (UAS)

UAS is a study that is supported by the Social Security Administration (SSA) and the National Institute on Aging (NIA). It has since 2014 collected more than 230 online surveys on various topics, ranging from cognitive abilities, environment, consumer behavior and politics to name but a few, for which the data is publicly available. [17] The UAS uses address-based sampling with sequential sample batching, where addresses were drawn from the Computerized Delivery Sequence (CDS) file created by the U.S. Postal Service. Annual attrition rates are modest (on the order of 8-9% per year).

The provision of tablets and free Internet to households without Internet access solves a coverage problem faced by convenience Internet panels. Respondents without prior Internet access have a very different demographic and socio-economic profile than respondents with Internet access such as they are more likely to have low incomes and education, to be non-white, lower health status and older (70+ years). However, among the non-Internet households, the probability of signing up for UAS is not related to these background characteristics, expect for age. One may interpret these findings as an indication that with respect to the demographics studied, the recruited non-Internet households are representative of the part of the population without Internet (except possibly with respect to age). ^[17]^

### 2. The PhQ-4 and Covid-19-related questions

The PhQ-4 is composed of the following four questions: Over the last two weeks, how often have you been bothered by any of the following problems?

- Feeling nervous, anxious, or on edge
- Not being able to stop or control worrying
- Feeling down, depressed, or hopeless
- Little interest or pleasure in doing things

Possible answers to these questions are: 0 Not at all, 1 Several days, 2 More than half the days, 3 Nearly every day. We added up the answers to these four questions and computed a PhQ-4 score, ranging from 0 to 12. We followed Kroenke et al. ^[21]^ and categorized respondents as having no depression/anxiety symptoms if their score equaled to 0, 1 or 2, mild depression/anxiety symptoms if their score ranged from 3 to 5, moderate depression/anxiety symptoms if their score ranged from 6 to 8 and severe depression/anxiety symptoms if their score ranged from 9 or above.

The question on the probability of being infected with the virus was phrased as: “On a scale of 0 to 100 percent, what is the chance that you will get the coronavirus in the next three months? If you’re not sure, please give your best guess.” The question on the probability of dying if infected was phrased as: “If you do get the coronavirus, what is the percent chance you will die from it? If you’re not sure, please give your best guess.” Finally, the question about the probability of running out of money because of the Covid-19 pandemic was: “The coronavirus may cause economic challenges for some people regardless of whether they are actually infected. What is the percent chance you will run out of money because of the coronavirus in the next three months?”

### 3. γ-standardization of the continuous variables

The coefficient of standardized continuous variable can be interpreted as the effect of a one standard deviation increase in that variable on the outcome of interest. On the other hand, the coefficient associated to a dichotomous variable corresponds to a marginal increase in that variable from 0 to 1, which is usually larger in magnitude than a standard deviation increase in the continuous variable. Gelman ^[39]^ therefore suggests to rescale the continuous variable by more than just their standard deviation so that the marginal effect of the continuous variables can be more meaningfully compared to the marginal increase in the dichotomous variable.

In essence, following the notation in the main text, Gelman’s ^[39]^ idea is to choose γ in such a way that the γ standard deviation increase in the continuous variables correspond to the marginal increase in the dichotomous variable from 0 to 1. One can therefore use the dichotomous variable characteristic as a benchmark for rescaling the continuous variables. Gelman ^[39]^ suggests to use γ = 2, which works well when the mean of the dichotomous variable is close or equal to 0.5. Indeed, if the mean of a dichotomous variable is equal to 0.5, then its standard deviation would be equal to 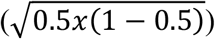. A two standard deviation increase in the continuous variable would therefore correspond to a marginal increase in the dichotomous variable from 0 to 1, as 1 is equal to two standard deviations of the dichotomous variables as well. In our study, because the mean of social distancing is equal to 0.655, we choose *γ* = 2.103. Indeed, 2.103times the standard deviation of social distancing equal to 1 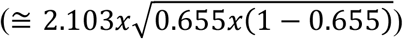 so that a *γ* × standard deviation increase in the continuous variables corresponds to an increase in the dichotomous variable from 0 to 1. The coefficients associated with the transformed continuous variables can then be meaningfully compared to the coefficient associated with the dichotomous variable.

**Table A1:** Descriptive statistics - additional variables

|  | Pop. mean | Pop. std. deviation |
| --- | --- | --- |
| Male | 0.489 | 0.500 |
| Age | 48.429 | 16.583 |
| Less than high school | 0.086 | 0.280 |
| High school graduate or GED | 0.297 | 0.457 |
| Some college (no degree) | 0.277 | 0.447 |
| College graduate and above | 0.341 | 0.474 |
| White | 0.772 | 0.420 |
| Black | 0.126 | 0.332 |
| Other races | 0.102 | 0.302 |
| Currently married | 0.557 | 0.497 |
| Hardware (internet-connected tablet) provided | 0.044 | 0.205 |

*Notes:* Source of Data: “Understand America Study” (UAS), survey 230, collected between March 10 and March 31, 2020. We use sample weights to make the survey representative of the U.S. population aged 18 and older. “Pop.” is short for population and “std.” is short for standard.

**Table A2:**
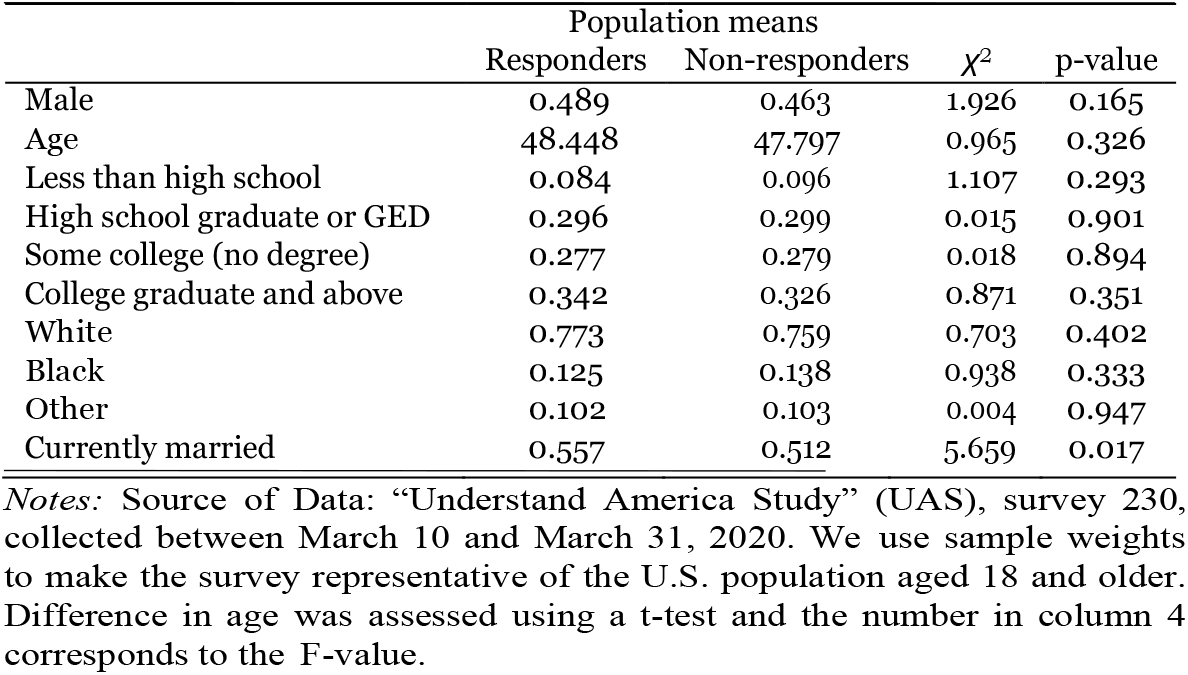
Differences between UAS responders and non-responders

| | Population means | | $\chi^2$ | p-value |
| --- | --- | --- | --- | --- |
|  | Responders | Non-responders |  |  |
| Male | 0.489 | 0.463 | 1.926 | 0.165 |
| Age | 48.448 | 47.797 | 0.965 | 0.326 |
| Less than high school | 0.084 | 0.096 | 1.107 | 0.293 |
| High school graduate or GED | 0.296 | 0.299 | 0.015 | 0.901 |
| Some college (no degree) | 0.277 | 0.279 | 0.018 | 0.894 |
| College graduate and above | 0.342 | 0.326 | 0.871 | 0.351 |
| White | 0.773 | 0.759 | 0.703 | 0.402 |
| Black | 0.125 | 0.138 | 0.938 | 0.333 |
| Other | 0.102 | 0.103 | 0.004 | 0.947 |
| Currently married | 0.557 | 0.512 | 5.659 | 0.017 |

*Notes:* Source of Data: “Understand America Study” (UAS), survey 230, collected between March 10 and March 31, 2020. We use sample weights to make the survey representative of the U.S. population aged 18 and older. Difference in age was assessed using a t-test and the number in column 4 corresponds to the F-value.

**Table A3:** Associations between PhQ-4 and raising number of Covid-19 cases and deaths during March 10-31, 2020

|  | PhQ-4 | PhQ-4<br>Female | PhQ-4<br>Male | PhQ-4<br>No college | PhQ-4<br>College | PhQ-4<br>White | PhQ-4<br>Non-white |
| --- | --- | --- | --- | --- | --- | --- | --- |
|  | (1) | (2) | (3) | (4) | (5) | (6) | (7) |
| **Number of cases in US (log)** | 0.090*** | 0.074*** | 0.113*** | 0.070** | 0.137*** | 0.093*** | 0.082 |
|  | [0.044,0.136] | [0.027,0.122] | [0.039,0.187] | [0.014,0.126] | [0.080,0.195] | [0.046,0.141] | [-0.017,0.182] |
| Observations | 6514 | 3790 | 2724 | 3844 | 2670 | 5084 | 1430 |
| **Number of deaths in US (log)** | 0.104*** | 0.086*** | 0.131*** | 0.080** | 0.157*** | 0.107*** | 0.099* |
|  | [0.051,0.157] | [0.031,0.141] | [0.044,0.218] | [0.015,0.146] | [0.090,0.224] | [0.050,0.163] | [-0.017,0.215] |
| Observations | 6514 | 3790 | 2724 | 3844 | 2670 | 5084 | 1430 |

*Notes:* Results of weighted ordered probit regressions with cluster robust confidence intervals at the state level in brackets, \* $p < 0.1$ , \*\* $p < 0.05$ , \*\*\* $p < 0.01$ . The number of cases and deaths in the US is transformed using log. Specifications include state fixed effects. The list of control variable includes: sex, age and age<sup>2</sup>, educational level (binary variable for each category), race and whether the respondent was married at the time of the interview. We control for depression level at baseline, along with the year and month when this measure was collected. We use sample weights to make the survey representative of the U.S. population aged 18 and older. Data come from “Understanding America Study” (UAS) collected between March 10 and March 31, 2020.

**Figure A1:**
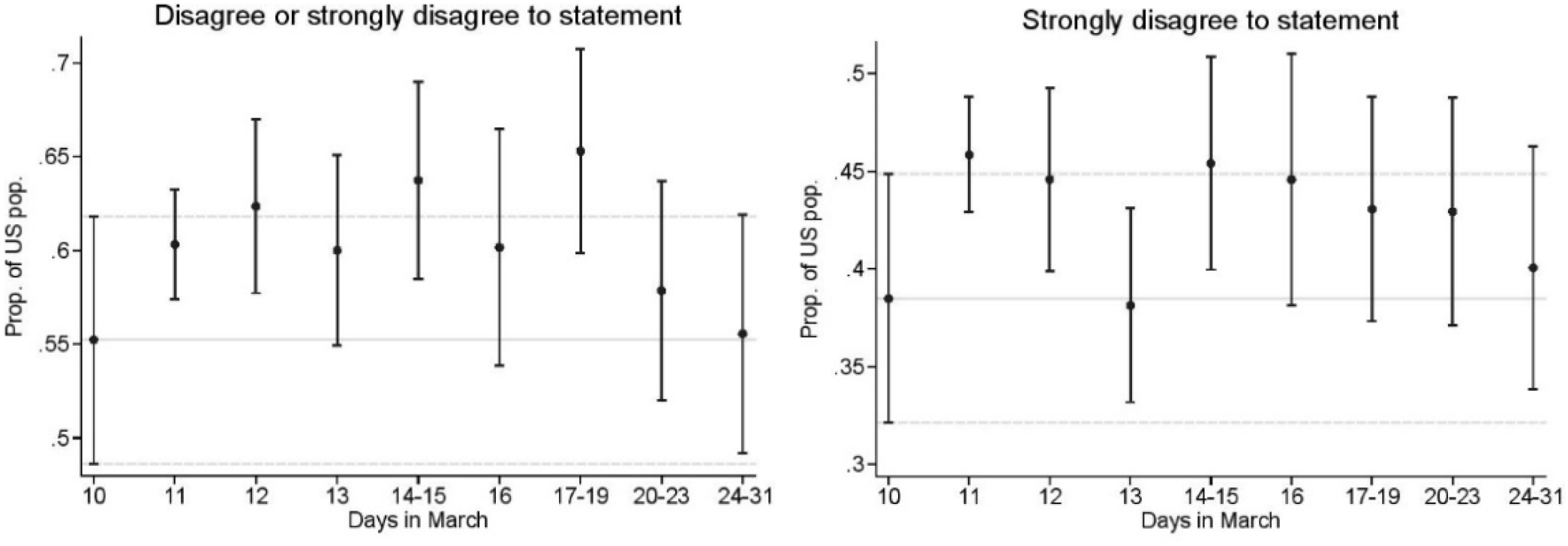
Weighted means and proportions of baseline depression between March 10-31 *Notes*: Source of Data: “Understanding America Study” (UAS), survey 230, collected between March 10 and March 31, 2020. Plots shows weighted means, along with 95% confidence intervals, of Phq-4 score on the left and weighted proportions of the US population that has at least some depressive/anxiety symptoms on the right. We use post-stratification weights so that the weighted means are representative of the US population for *each* particular time period on the *x axis*. Baseline depression level is defined based on the question: “I see myself as someone who is depressed, blue”. The plot on the left shows the proportion of the US population that strongly disagrees to this statement while the one of the right shows the proportion did strongly disagree or disagree to the statement. This question was asked in UAS survey 121, fielded between Jan 2018 and March 2020.

**Table A4:** Associations with Covid-19 cases and related number of deaths and mental health score (PhQ-4) - Average marginal effects

|  | PhQ-4 categories |  |  |  |
| --- | --- | --- | --- | --- |
|  | None | Mild | Moderate | Severe |
| Number of cases in US (log) | -0.026***<br>[-0.040,-0.013] | 0.012***<br>[0.006,0.018] | 0.007***<br>[0.004,0.010] | 0.007***<br>[0.003,0.012] |
| Number of deaths in US (log) | -0.031***<br>[-0.046,-0.015] | 0.014***<br>[0.007,0.021] | 0.008***<br>[0.004,0.012] | 0.009***<br>[0.004,0.013] |

*Notes:* Average marginal effects resulting from weighted ordered probit regressions with robust confidence intervals clustered at the state level in brackets, \* $p < 0.1$ , \*\* $p < 0.05$ , \*\*\* $p < 0.01$ . The number of cases and deaths in the US is transformed using log. Specifications include state fixed effects. The list of control variable includes: sex, age and age<sup>2</sup>, educational level (binary variable for each category), race and whether the respondent was married at the time of the interview. We control for depression level at baseline, along with the year and month when this measure was collected. We use sample weights to make the survey representative of the U.S. population aged 18 and older. Data come from “Understanding America Study” (UAS) collected between March 10 and March 31, 2020.

**Table A5:**
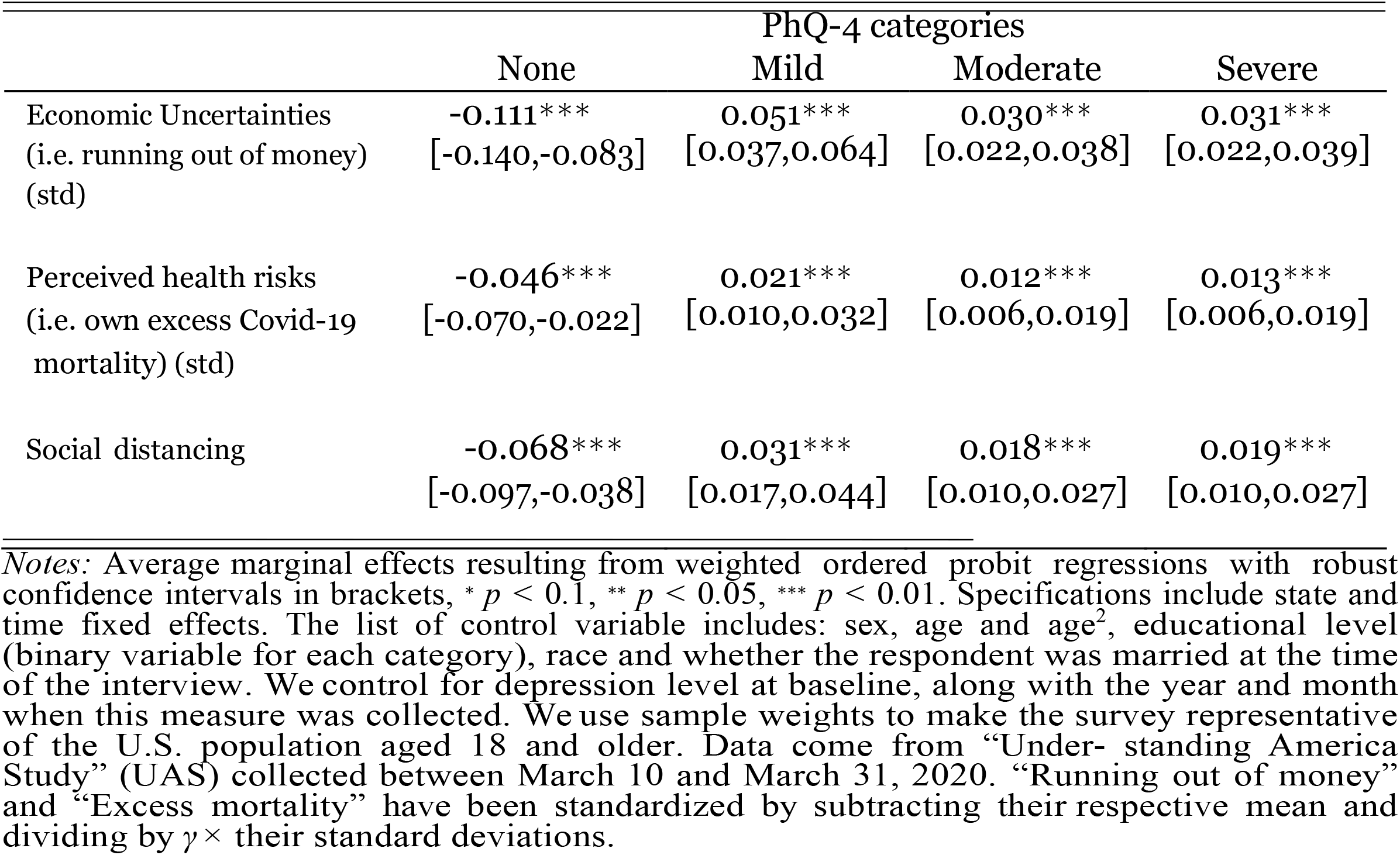
Associations with mental health score (PhQ-4) - Average marginal effects

**Table A6:** Associations with mental health score (PhQ-4) with our three channels, interacting them with three sociodemographic groups

|  | PhQ-4<br>Gender<br>(1) | PhQ-4<br>Education<br>(2) | PhQ-4<br>Race<br>(3) |
| --- | --- | --- | --- |
| Economic Uncertainties (i.e. running out of money) (std) | 0.311***<br>[0.186,0.435] | 0.416***<br>[0.296,0.535] | 0.367***<br>[0.256,0.478] |
| × Male | 0.230**<br>[0.035,0.425] |  |  |
| × College graduate |  | -0.045<br>[-0.279,0.189] |  |
| × Non-white |  |  | 0.099<br>[-0.136,0.334] |
| Perceived health risks (i.e. own excess Covid-19 mortality) (std) | 0.222***<br>[0.120,0.325] | 0.165***<br>[0.064,0.266] | 0.217***<br>[0.121,0.312] |
| × Male | -0.150*<br>[-0.323,0.023] |  |  |
| × College graduate |  | -0.001<br>[-0.177,0.174] |  |
| × Non-white |  |  | -0.224**<br>[-0.437,-0.010] |
| Social distancing | 0.212***<br>[0.078,0.346] | 0.202***<br>[0.070,0.334] | 0.237***<br>[0.125,0.350] |
| × Male | 0.055<br>[-0.151,0.262] |  |  |
| × College graduate |  | 0.136<br>[-0.067,0.339] |  |
| × Non-white |  |  | 0.027<br>[-0.258,0.312] |
| Observations | 6368 | 6368 | 6368 |

*Notes:* Results of weighted ordered probit regressions with robust confidence intervals in brackets, \* $p < 0.1$ , \*\* $p < 0.05$ , \*\*\* $p < 0.01$ . Specifications include state and time fixed effects. The list of control variable includes: sex, age and age<sup>2</sup>, educational level (binary variable for each category), race and whether the respondent was married at the time of the interview. We control for depression level at baseline, along with the year and month when this measure was collected. We use sample weights to make the survey representative of the U.S. population aged 18 and older. Data come from “Understanding America Study” (UAS) collected between March 10 and March 31, 2020. “Running out of money” and “Excess mortality” have been standardized by subtracting their respective mean and dividing by $\gamma \times$ their standard deviations.

## Data Availability

Undestanding America Data (UAS) used in this manuscript are publicly avaiable at: https://uasdata.usc.edu/index.php

